# Building Blocks for Integrating Image Analysis Algorithms into a Clinical Workflow

**DOI:** 10.1101/2020.03.06.20027367

**Authors:** Krishna Juluru, Hao-Hsin Shih, Pierre Elnajjar, Amin El-Rowmeim, Josef Fox, Eliot Siegel, Krishna Nand Keshava Murthy

## Abstract

**Purpose:** Starting from a broad-based needs assessment and utilizing an image analysis algorithm (IAA) developed at our institution, the purpose of this study was to define generalizable building blocks necessary for the integration of any IAA into a clinical practice.

**Methods:** An IAA was developed in our institution to process lymphoscintigraphy exams. A team of radiologists defined a set of building blocks for integration of this IAA into clinical workflow. The building blocks served the following roles: (1) Timely delivery of images to the IAA, (2) quality control, (3) IAA results processing, (4) results presentation & delivery, (5) IAA error correction, (6) system performance monitoring, and (7) active learning. Utilizing these modules, the lymphoscintigraphy IAA was integrated into the clinical workflow at our institution. System performance was tested over a 1 month period, including assessment of number of exams processed and delivered, and error rates and corrections.

**Results:** From June 26-July 27, 2019, the building blocks were used to integrate IAA results from 132 lymphoscintigraphy exams into the clinical workflow, representing 100% of the exams performed during the time period. The system enabled radiologists to correct 21 of the IAA results. All results and corrections were successfully stored in a database. A dashboard allowed the development team to monitor system performance in real-time.

**Conclusions:** We describe seven building blocks that optimize the integration of IAAs into clinical workflow. The implementation of these building blocks in this study can be used to inform development of more robust, standards-based solutions.

## INTRODUCTION

Artificial intelligence (AI), machine learning, and deep learning applications are increasingly being used in diagnostic imaging (1). Several of these applications have been described in the literature and can be divided broadly into two categories: first, those pertaining to logistical workflow, such as order scheduling, patient screening, and other operational analytics; and second, those pertaining to the acquired imaging data itself, such as automated detection of findings or features, automated interpretation of findings, and image postprocessing (2). The latter category of varied applications can be generally named ‘Image Analysis Algorithms (IAA); the availability and scope of these IAAs have grown in recent years due to the growth in computer science expertise and computational power.

At our institution, the Radiology Informatics team built an IAA to process images from a type of examination performed in our Nuclear Medicine department known as ‘Lymphoscintigraphy’. The current paper focuses on the incorporation of any IAA into the clinical workflow to enable the optimal use of its output. Standards-based solution and general guidance in addressing the challenges of clinical workflow integration is limited in current literature.

Starting from a broad-based needs assessment and using the Lymphoscintigraphy IAA as a use-case, the purpose of this study was to define generalizable building blocks necessary for the integration of any IAA output into a clinical practice. The goal is for these building blocks and concepts to facilitate the integration of other IAAs into clinical workflows.

## MATERIALS / METHODS

### UNDERSTANDING THE GENERAL WORKFLOW AND PARTICULAR USE-CASE

The lymphoscintigraphy exam is a long-established procedure that is performed to identify sentinel lymph nodes in patients with invasive breast cancer, potentially increasing the yield of axillary lymph node dissections and the accuracy of staging (3). It is performed after breast cancer has been diagnosed using other techniques. In the routine workflow at our institution, a surgical oncologist orders the lymphoscintigraphy exam preoperatively on a patient with known breast cancer, specifying the right, left, or bilateral breasts. Within the Nuclear Medicine department, staff inject a radioactive substance into the patient’s breast(s), which then travels through the lymphatic system to a subset of the lymph nodes in the nearby axilla that are most likely to be locations of regional spread of cancer. These nodes are referred to as “sentinel nodes” (SLN). A technologist acquires images of the breasts and axilla using a gamma camera (Figure 1); the images are sent to the Picture Archiving and Communication System (PACS). A resident radiologist opens the exam in PACS and performs routine quality checks, including ensuring that the site of injection matches the site requested by the surgical oncologist. He then generates a preliminary report using a dictation software, describing the site of injection, the presence of radiotracer accumulation (if any) in the axillae, and the number of these sites. An attending radiologist later reviews the report and signs it. The final report is sent to the institution electronic medical record, available for viewing by the clinical team. At the time of operation, the surgeon identifies these SLNs using a probe that detects radioactivity, and removes them for pathological diagnosis.

**Figure 1.**
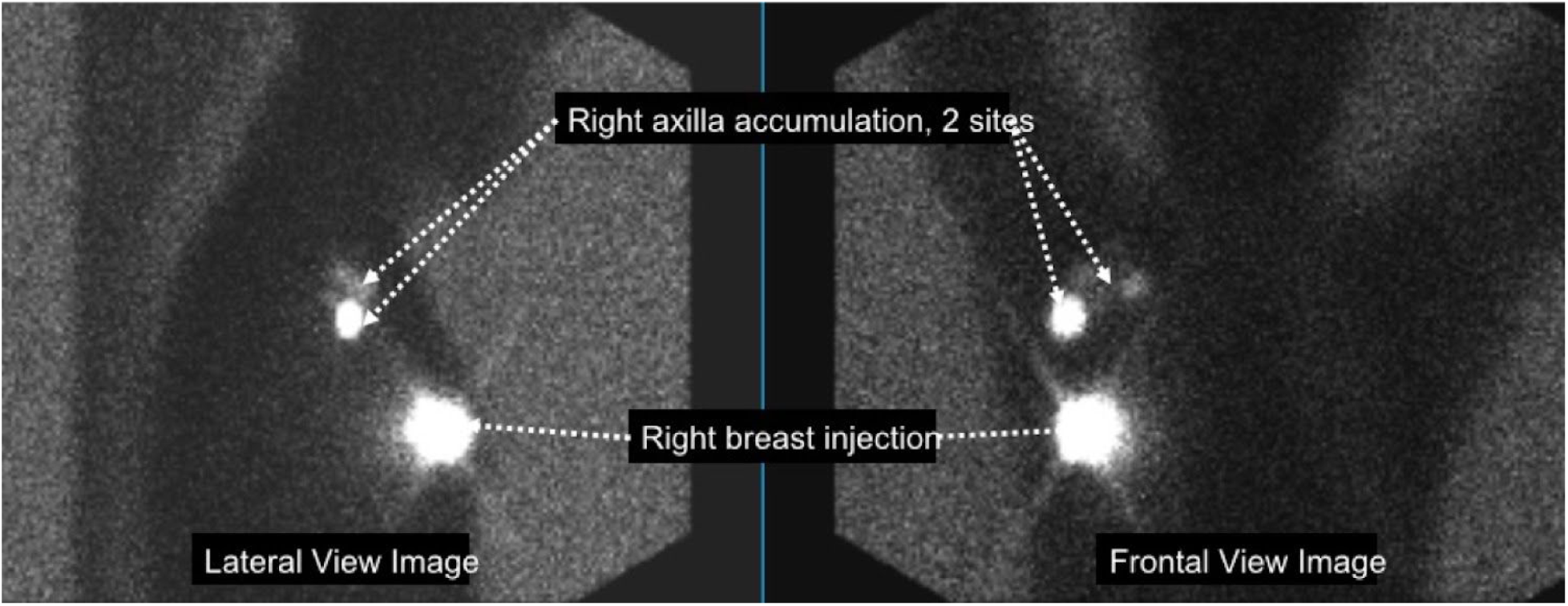
Images obtained from a lymphoscintigraphy exam. Following injection of a radioactive tracer into one or both breasts, images are obtained with a gamma camera in anterior and lateral projections. Bright areas are sites of injection or radiotracer accumulation in lymph nodes, where breast cancer may have spread. In the current example, there was a right breast injection, with two sites of accumulation in the right axilla.

The Radiology Informatics team at our institution built an IAA that processes images from these lymphoscintigraphy exams and generates the following outputs (Table 1): (1) observed sites of injection (right breast only, left breast only, bilateral breasts), (2) probability of radiotracer accumulation in the axillae (probability scores for none, right, left, or bilateral axillae),(3)number of right axillary lymph nodes (integer), and (4) number of left axillary lymph nodes (integer).

**Table 1.** Results database fields storing lymphoscintigraphy data:

| Field | Source | Possible values |
| --- | --- | --- |
| Ordered exam accession number | PACS / RIS Monitor | Integer |
| Ordered exam name | PACS / RIS Monitor | Text |
| Injection site | IAA | Right, left, bilateral |
| Probability of radiotracer accumulation in no axilla* | IAA | Value between 0 and 1 |
| Probability of radiotracer accumulation in right axilla* | IAA | Value between 0 and 1 |
| Probability of radiotracer accumulation in left axilla* | IAA | Value between 0 and 1 |
| Probability of radiotracer accumulation in bilateral axillae* | IAA | Value between 0 and 1 |
| Number of right axillary lymph nodes | IAA | Integer |
| Number of left axillary lymph nodes | IAA | Integer |
| Axillary sites | Reporting Module<br>(calculated result from<br>probability scores) | None, right, left, bilateral |
| Correction injection site | Error correction module | Right, left, bilateral |
| Corrected axillary sites | Error correction module | None, right, left, bilateral |
| Corrected number of right axillary lymph nodes | Error correction module | Integer |
| Corrected number of left axillary lymph nodes | Error correction module | Integer |

Abbreviations: IAA, image analysis algorithm
\* Sum of all probabilities = 1

### NEEDS ASSESSMENT

The Radiology Informatics team met with key physicians to assess the general needs of a system that integrates an IAA into clinical workflow, with the lymphoscintigraphy IAA as a use-case. The following needs were identified for a system integrating IAA results into a clinical workflow.

1. System should begin image analysis soon after completion of image acquisition
2. System should identify and report exam quality problems
3. System should generate exam preliminary reports and send them to the dictation system for radiologist review and signature
4. System should allow users to correct IAA results
5. System should use corrected results to retrain IAA
6. IAA results, both initial and corrected, should be accessible on a dashboard to allow users to monitor system performance

### BUILDING BLOCKS / MODULES NECESSARY TO MEET NEEDS

Using the needs assessment as a foundation, we developed seven generalizable building blocks for the integration of an IAA into any clinical workflow, with the lymphoscintigraphy IAA as a use-case (Figure 2). All databases were implemented on SQL Server 2012 SP4 (Microsoft Corporation, Redmond, VA). To enable timely and autonomous analysis, all software components were implemented as operating system services that ran continually in the background, monitoring for changes. The IAA itself was implemented as a web service, where the various modules communicated via Application Programming Interface (API) calls. A web portal was built to gather all user feedback / input. All software ran on Microsoft Windows Server 2012 R2. All programming was done using C# (version 5.0) and Python (version 3.5) languages.

**Figure 2.**
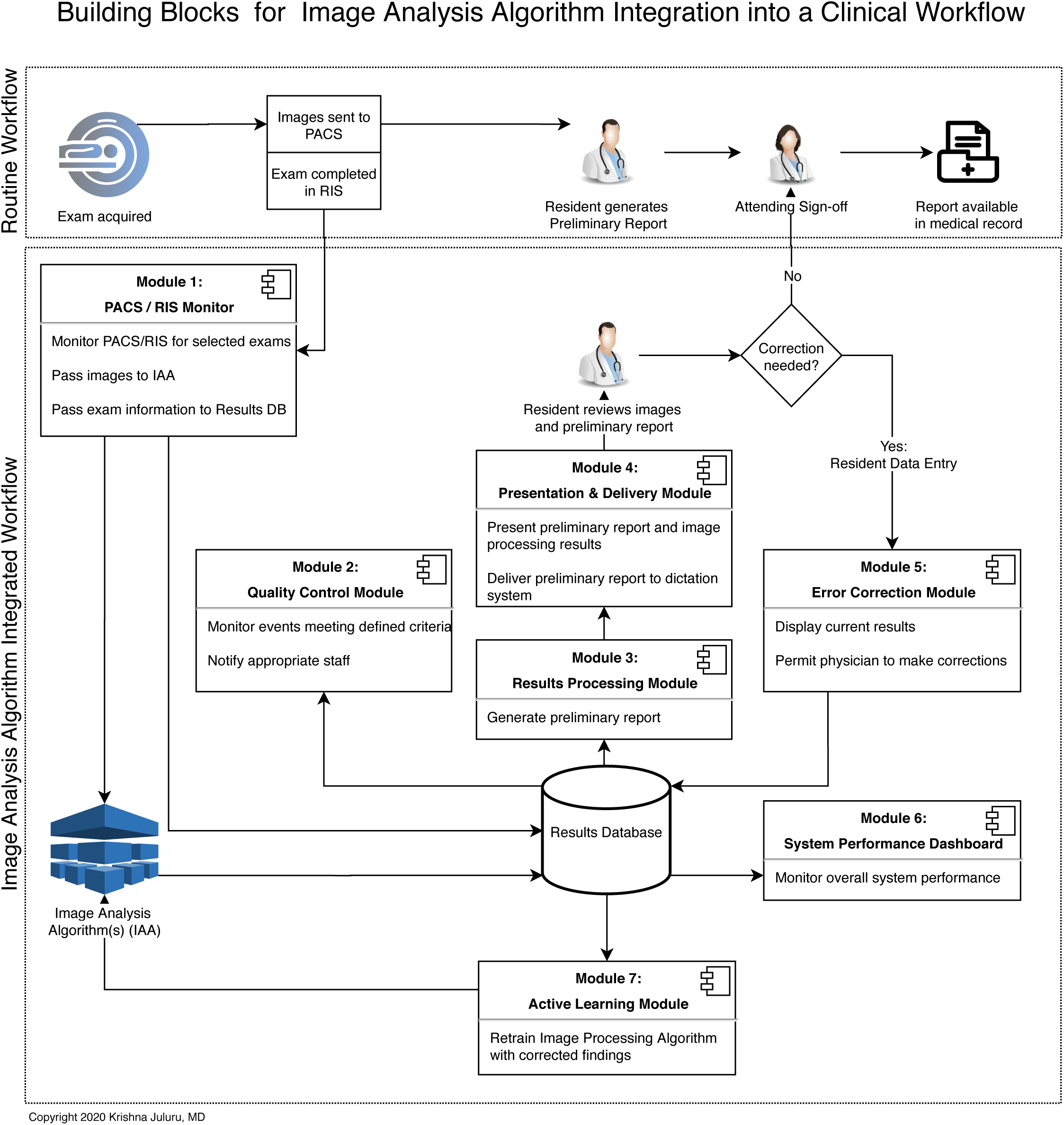
Routine clinical workflow is compared with a workflow that includes an image analysis algorithm that performs a preliminary image analysis. Seven building blocks are described that are necessary for integration into a clinical environment.

### PACS / Radiology Information System (RIS) Monitor Module

The PACS / RIS Monitor Module is a building block that allows the system configuration team to specify a list of exams to monitor in PACS or RIS. When the specified exams are completed, the monitor extracts the images from PACS and delivers them to the correct IAA, thereby meeting Needs Assessment #1. The module also sends specified exam information to the database.

There are three distinct orders for lymphoscintigraphy exams in our institution. A physician may enter a request for (1) Lymph scan right breast, (2) Lymph scan left breast, or (3) Lymph scan bilateral breasts, each specifying the requested site(s) of injection of the radioisotope, corresponding to the site(s) of known breast cancer. We configured our PACS / RIS Monitor Module to monitor for these exams. Once the technologist completes an exam in RIS, the module would send a copy of the images from PACS to the Lymphoscintigraphy IAA.

### Quality Control Module

The Quality Control Module is a building block that provides a means for the system to identify and then notify the clinical team of issues arising in the workflow or in the quality of imaging, thereby meeting Needs Assessment #2. Notifications of issues can take several forms, such as emails or pages. In our implementation of this module, we incorporated an open-source ticketing system that was previously implemented in a quality improvement effort (4), whereby a ticket would be filed into the ticketing system through an API call whenever an issue is detected, and where the rules engine within the ticketing system can be configured to send an email to a quality control team member appropriate to the particular examination with the issue.

For the Lymphoscintigraphy project, we desired to identify the unlikely event when the wrong breast has been injected by finding a mismatch between the order placed and the site of injection detected by the IAA (Table 2). Recall that the exam order allowed the surgical oncologist to specify the site of injection as right, left, or bilateral breasts. If the IAA result of observed site of injection did not match the site requested by the surgical oncologist, the Quality Control Module was set to activate.

**Table 2.**
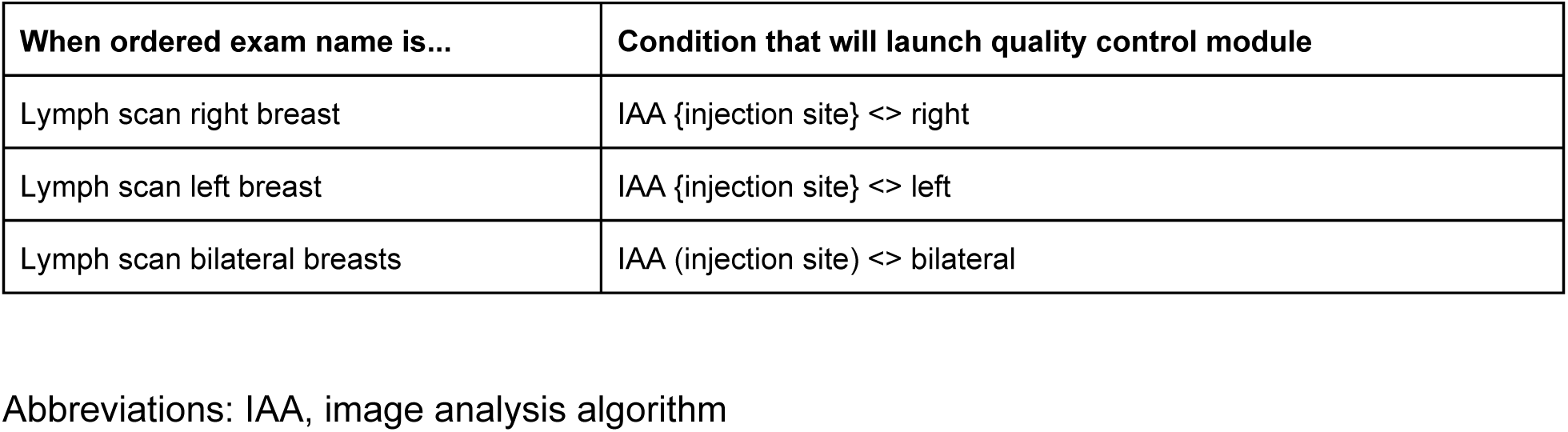
Quality Control Module rules established for lymphoscintigraphy project.

### Results Database

The Results Database is a building block that stores information about the exam from the PACS / RIS monitor, the results from the IAA, and corrections from the Error Correction Module. It serves as the source of data for the Results Processing Module, System Performance Dashboard, and Active Learning Module. Our database was implemented on SQL Server 2012 SP4, which was available through an institutional site license.

For the Lymphoscintigraphy project, the fields, information sources, and possible values of items in the Results Database are summarized in Table 1.

### Results Processing Module

The Results Processing Module is a building block that generates a preliminary report based on the available data in the Results Database. The preliminary report may be a simple incorporation of IAA results into a descriptive, human-friendly narrative. A more sophisticated implementation is one where this module uses database information to make calculations that are used to adjust the language in the final report.

For the Lymphoscintigraphy project, the Results Processing Module compiled results for the ‘FINDINGS’ and ‘IMPRESSION’ sections of the preliminary report, including the observed sites of injection, sites of axillary radiotracer accumulation, and number of lymph nodes in each axilla (Table 3). Recall that this IAA does not provide sites of axillary radiotracer accumulation as a single result, but rather provides probabilities for ‘no axilla’, ‘right axilla’, ‘left axilla’, or ‘bilateral axilla’ (Table 1). Our results processing module was configured in this project, therefore, to calculate the site of highest probability to use in the preliminary report.

**Table 3.** Sample Lymphoscintigraphy Report. Findings and Impression are automatically generated by the Results Processing Module and delivered to a custom field in the dictation system. The custom field is a part of a template that is set to automatically launch for the lymphoscintigraphy exams. The net effect was one in which at the moment the radiologist opened a lymphoscintigraphy exam for dictation, he/she would see the entire report ready for review and signature. The radiologist’s next step was to ensure agreement between the imaging examination in PACS and the report. If this agreement existed, the radiologist need only to sign the report.

| Final report text sent to medical record | Comment |
| --- | --- |
| **CLINICAL STATEMENT:** Breast cancer; referred for sentinel lymph node mapping. | Standard text, part of dictation system default template. |
| **TECHNIQUE:**<br>Radiopharmaceutical: Right breast 0.102 mCi Tc-99m sulfur colloid (unfiltered), 6 o'clock subareolar<br>Field of view: Anterior and lateral images of the chest with transmission images for anatomic localization.<br>**COMPARISON:** None<br>**CORRELATION:** None | Radiopharmaceutical dose (highlighted) is extracted from PACS technologist notes and automatically inserted into dictation system custom field by Presentation & Delivery Module.<br>Remainder of text is part of dictation system default template. |
| **FINDINGS:**<br>Radiotracer accumulation is seen in the right breast.<br>Sites of radiotracer accumulation outside of the breast:<br>- Right axilla: 2<br>**IMPRESSION:**<br>Sentinel lymph nodes in the right axilla. | All text in this section (highlighted) is generated by the Results Processing Module. In this example, results database fields for probabilities of radiotracer accumulation in no axilla, right, left, and bilateral axilla were 0.08, 0.88, 0.02, 0.02, respectively. The Results Processing Module selected the highest probability to include in the report.<br>The Presentation & Delivery Module delivered the report into the dictation system custom field. |

### Presentation & Delivery Module

The Presentation & Delivery Module is a building block that takes the output of the Results Processing Module and delivers it into the dictation system for review by the radiologist, thereby meeting Needs Assessment #3. We combined the steps of presentation and delivery into a single step to meet the needs of this project, although they can be split into separate steps when necessary, as reviewed in the Discussion section. Our dictation system (PowerScribe® 360 | Reporting v4.0 SP1, build 7.0.111.5, Nuance Communications, Inc, Burlington, MA) allowed administrators to create user-defined ‘custom fields’. These custom fields could be inserted into reporting templates known as ‘AutoTexts’ that could be set to launch by default for specific examinations. Our code leveraged an API available within our dictation system that allows the delivery of any text into the custom field.

For the Lymphoscintigraphy project, we created a custom field in the dictations sytem called “AI Result” that was included as part of an AutoText called, “Lymphoscintigraphy”. The Presentation & Delivery Module would send the ‘FINDINGS’ and ‘IMPRESSION’ text generated by the Results Processing Module to this custom field. The AutoText was set to launch by default when any lymphoscintigraphy exam is opened by a radiologist. The net effect was one in which when the radiologist opened a lymphoscintigraphy exam for dictation he/she would see the entire report ready for review and signature. The radiologist’s next step was to ensure agreement between the imaging examination in PACS and the report. If this agreement existed, the radiologist need only to sign the report.

### Error Correction Module

The Error Correction Module is a building block that allows the radiologist to correct errors in the IAA results as observed in the report, thereby satisfying Needs Assessment #4. Although the radiologist can correct errors simply by editing the text of the report in the dictation system, this approach would fail to discreetly capture the changes made to support active learning, the next module. Thus, the Error Correction module is considered an essential building block. Our Error Correction Module utilized features in our PACS (Centricity RA1000, GE Healthcare, Chicago, IL) that permitted an administrator to assign a button, available for all users, that performed context passing of data of the currently-viewed exam (eg. current user, exam accession number) to a web application, a methodology previously described for quality reporting (4). The web application would display current IAA results from the database, with options to correct those results, and then send the corrected data back to the database. Subsequently, the Results Processing and Presentation & Delivery Modules would be automatically re-initiated, thereby generating a new report in the dictation system for radiologist re-review.

For the Lymphoscintigraphy project, the Error Correction Module could be launched by the resident by selecting the pre-configured button on our PACS that passed the patient context to a web application. The webpage would display a form showing the database fields and current processing results (Figure 3). The radiologist would be able to make corrections to these fields as needed, and the values would be returned to a new set of fields in the same record of the database (Table 1). Updates to the database would re-trigger the Results Processing and Presentation & Delivery Modules so that an updated report is delivered to the dictation system for Radiologist re-review.

**Figure 3.**
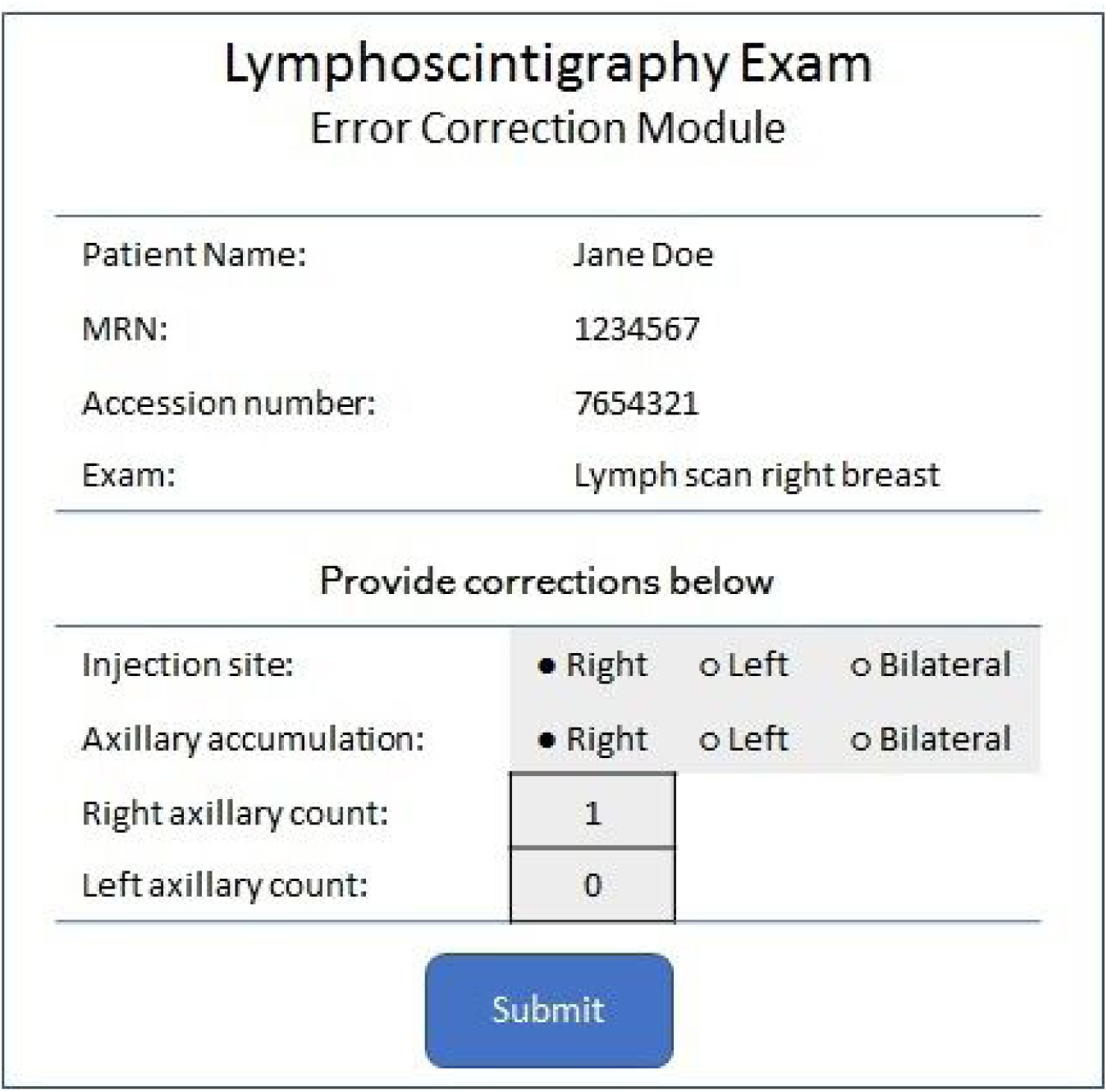
Error Correction Module-generated form that allows radiologists to provide corrections to report information. The Radiologist launches the form from PACS. After clicking ‘Submit’, the data is recorded in the database, and the preliminary report is recreated and delivered to the dictation system by other modules in the system.

### Active Learning Module

The Active Learning Module is the same as, or a variation of, the original training toolset for creating the IAA. Together with the Error Correction module, it meets Needs Assessment #5. It is specific to the needs of the individual application, including its unique inputs and outputs.

Details on the design and development of the Lymphoscintigraphy IAA are to be described in a separate paper.

### Dashboarding tool

There are a variety of dashboarding and business intelligence tools on the market that enable data visualization (5). Our institution has an enterprise-level subscription to one such tool known as Tableau (Tableau Server Version: 2019.1.1 (20191.19.0215.0259) 64-bit Windows, Tableau Software, Inc, Mountain View, CA) that has been applied to healthcare data visualization challenges (6), thereby meeting Needs Assessment #6.

For the Lymphoscintigraphy project, we utilized Tableau to generate real-time metrics of data available in the results database, including quantifying discrepancies between IAA results and corrected results (Figure 4).

**Figure 4.**
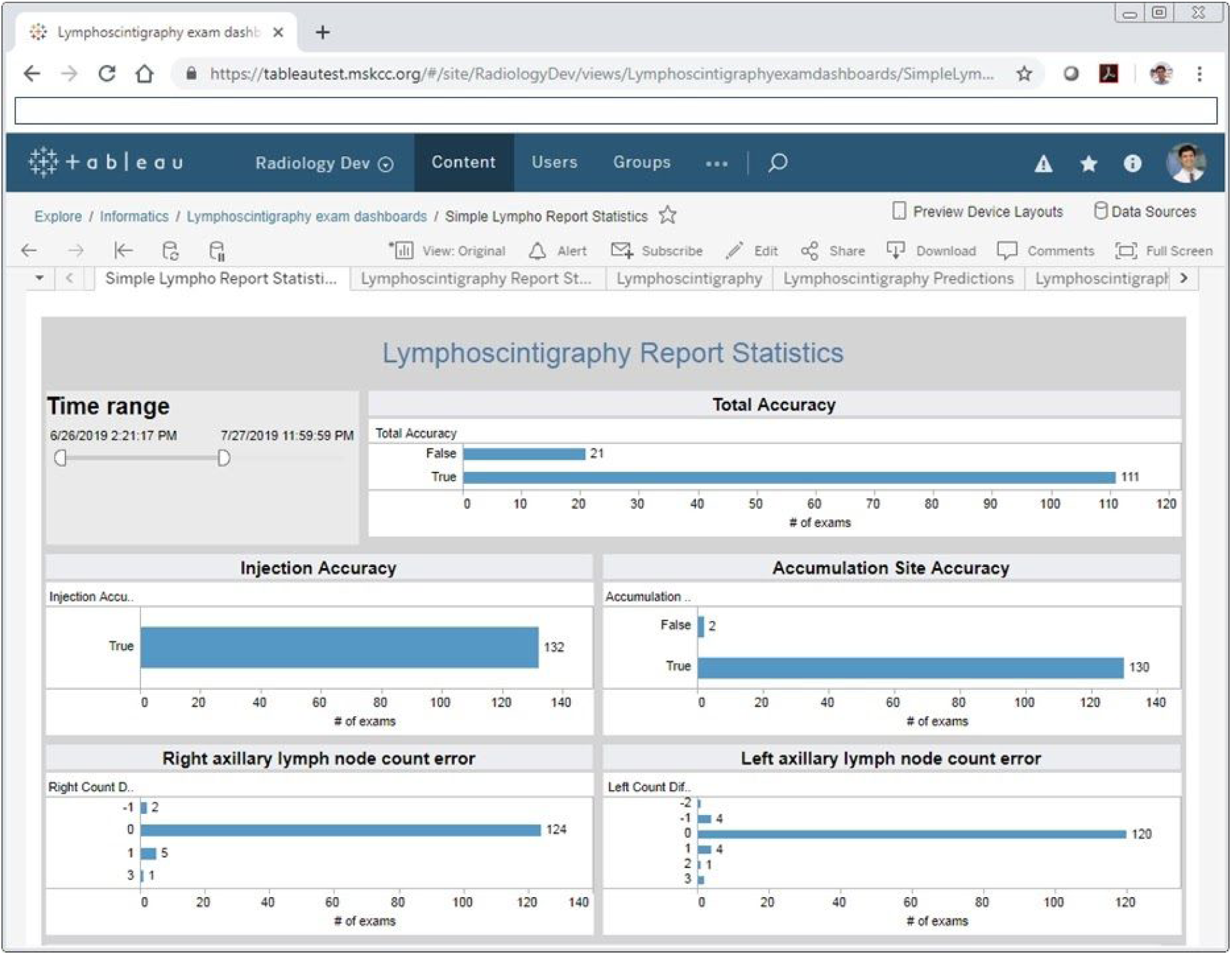
The dashboard specific for the lymphoscintigraphy image analysis algorithm, utilized as a use-case in this study, allows close monitoring of algorithm performance (Module 6 in Figure 2). The dashboard draws data in near real-time from the system database, and is accessible from a web-browser. Total accuracy is TRUE if no corrections were made to any field in the Results Database, otherwise FALSE. Injection accuracy is TRUE if no correction was made to the Injection Site field, otherwise FALSE. Axillary site accuracy is TRUE if no correction was made to the axillary site field, otherwise FALSE. Right axillary lymph node count error = (Corrected number of right axillary lymph nodes following radiologist correction) - (Number of right axillary lymph nodes originally identified by the IAA). Left axillary lymph node count error = (Corrected number of left axillary lymph nodes following radiologist correction) - (Number of left axillary lymph nodes originally identified by the IAA).

### DEPLOYMENT AND DATA ANALYSIS

Attending faculty radiologists and radiology residents were trained in using the system during June 2019 using retrospective data. On June 26, 2019, the system was turned on to process exams and deliver reports prospectively. Data were extracted for analysis on July 27, 2019. Algorithm and system performance were assessed using the following metrics obtained from the Results Database (Table 1):

Total accuracy is TRUE if no corrections were made to any field in the Results Database, otherwise FALSE. Injection accuracy is TRUE if no correction was made to the Injection Site field, otherwise FALSE. Axillary site accuracy is TRUE if no correction was made to the axillary site field, otherwise FALSE.

Right axillary lymph node count error = (Corrected number of right axillary lymph nodes following radiologist correction) - (Number of right axillary lymph nodes originally identified by the IAA).

Left axillary lymph node count error = (Corrected number of left axillary lymph nodes following radiologist correction) - (Number of left axillary lymph nodes originally identified by the IAA).

## RESULTS

System performance metrics were readily available on the Tableau Dashboard (Figure 4). In the one-month pilot period from June 26–July 27, 2019, the system processed 132 lymphoscintigraphy exams and delivered 132 reports to the dictation system for resident review. Of these 132 exams, overall report accuracy was TRUE for 111 exams (accuracy rate = 111/132 = 84%). Injection accuracy was TRUE for 132 exams (accuracy rate = 132/132 = 100%). Axillary accumulation site accuracy was TRUE for 130 exams (accuracy rate = 130/132 = 98%).

Right axillary lymph node count error was ‘0’ for 124 exams. In 6 exams, the right axillary count error was a positive value of either ‘1’ or ‘3’, meaning that the IAA undercounted the number of right axillary lymph nodes. In an additional 2 exams, the right axillary count error was ‘-1’, meaning that the IAA overcounted the number of right axillary lymph nodes.

Left axillary lymph node count error was ‘0’ for 120 exams. In 7 exams, the left axillary count error was a positive value ranging from ‘1’ to ‘3’. In 5 exams, the left axillary count error was a negative value ranging from ‘-1’ to ‘-2’.

A query of our RIS records confirmed that in fact 132 lymphoscintigraphy exams were performed during this time period, proving that the PACS / RIS Monitor Module, Results Database, Results Processing Module, and Presentation & Delivery Module worked as expected. The residents were successfully able to correct 21 out of the 132 reports generated, confirming proper functioning of the Error Correction Module.

The Quality Control Module was never activated during the time period of this study. We confirmed that there were no issues that met the criteria for activation of this module.

## DISCUSSION

The purpose of this study was to understand, develop, and test the building blocks needed to incorporate the output of an image analysis algorithm (IAA) into a modern clinical workflow in an academic medical center. A needs assessment generated in conjunction with clinical physicians identified six key requirements for a solution that would deliver results to our radiologists quickly, automate certain quality control processes, allow our radiologists to correct IAA result errors, support the ability to retrain the IAA with new data, and permit a real-time environment to monitor system performance. This needs assessment subsequently informed the design and development of seven key building blocks to create a system that can be re-used and adapted to a variety of IAAs serving a variety of functions.

The building blocks described serve the following specific functions, and all are designed to work with minimal or no human intervention. The first building block, the PACS / RIS monitor, needs a one-time configuration to define which exams need to be processed. Subsequently, it monitors the PACS / RIS for these exams and delivers images to the IAA, allowing analysis to begin without delay. Future enhancements could provide the ability to define additional criteria such as referring physician and time of examination. Current standards that can be adapted to serve this function of the PACS / RIS monitor include the DICOM/IHE Instance Availability Notification (IAN) (7). DICOMweb standards can be used to support offsite image analysis, which would be limited by more traditional DICOM transfer methods (8).

The second building block, the Quality Control Module, provides a much-needed service that is difficult to provide in a current, high-volume radiology department, namely screening for procedure errors prior to a radiologist review. In typical outpatient settings, a patient leaves the imaging facility shortly after the completion of the examination, while a radiologist reviews the images some time afterward. Problems with image acquisition, if present, can be difficult to address or correct at this later time. There are benefits to patient safety, image quality, department logistics, and department finances if some problems are detected early and corrected promptly. In our lymphoscintigraphy use-case, the Quality Control Module was configured to compare the side of breast injection *ordered* with the side of breast injection *detected*. A mismatch could indicate that the referring physician communicated a change in site of injection without following through on a change in order or that a more serious event occurred where the wrong breast was injected. This would be a rare event and one that did not occur during the course of our study, but early detection could alert the staff so that they may repeat the procedure in the correct breast. These laterality errors have been previously described, and worst-case outcomes included wrong site surgery (9,10). Other studies have described automated algorithms for the detection of intravenous contrast in CT examinations (11). A Quality Control Module configured for this algorithm could identify differences between an order for a CT with contrast and the presence of contrast within the acquired images. A mismatch between what was ordered and what is present on the images could indicate a last-minute decision to not give contrast due to patient allergy or other contraindication. Alternatively, such a mismatch could also indicate a more serious scenario of iatrogenic contrast extravasation into nonvascular tissue, a condition requiring observation and treatment (12). As a final example, the Quality Control Module could be configured to identify specified high probability IAA results that could constitute critical results. A high probability of stroke on non-contrast head CT could trigger the Quality Control Module to alert a radiologist to prioritize review and reporting of that CT examination (13).

The third building block, the Results Processing Module, can be used to make decisions from IAA results in preparation for presentation to the radiologist. Processed results can take several forms. They can be a text/numbers intended for radiologist review or insertion into a report. Alternatively, they can be image overlays intended to draw attention to a particular region in a radiographic image. Some results may require no processing at all. For example, the IAA result of the aforementioned stroke detection algorithm (13) could include not only a probability score for likelihood of stroke but also an annotation that can be overlaid on the slice(s) where stroke is detected, for review and confirmation by the radiologist. Clunie provides an excellent review of available, standardized annotation formats (14). Annotation files can be stored in the results database or even in PACS, without further processing. Other IAA results may require further processing before presentation. In our lymphoscintigraphy use-case, we configured our Results Processing Module to perform the simple task of taking the IAA accumulation site result of highest probability (no axilla, right, left, or bilateral axilla) and generating a report that included that result only into the report (Table 3). This module can be applied more broadly, for instance, by combining IAA results with other data in the clinical record. For example, breast cancer risk has been shown to be correlated with clinical parameters of age, family history, race, and body mass index, and also with imaging parameters of breast density on mammography (15,16). IAAs that generate automated breast density values have been reported in the literature (15,16). A Results Processing Module can be used to combine clinical and imaging parameters to generate a combined risk score to be included in a radiology report.

The fourth building block, the Presentation & Delivery Module, is intended to deliver preliminary processed results to the radiologist for review. The presentation can take several forms and will need to be adjusted based on the output of the Results Processing Module. Several standards do currently exist that support results storage, such as AIM, DICOM SR, DICOM SEG, and DICOM Parametric Maps (17). Many, if not most PACS, however, do not support visualization of these data, necessitating other intermediary solutions to enable the workflow. In our use case, the result was text only, that we were able to deliver using a proprietary API available in our dictation system. Standards-based approaches to the delivery of this textual information could leverage technologies such as HL7 and FHIR DiagnosticReport (18).

The promise of machine learning algorithms, and one feature that distinguishes them from early rule-based CAD technologies, is their ability to be retrained on newly available data. These active learning paradigms will allow these algorithms to learn through smaller, more focused data that represent a wider breath of ground-truth labeled results (19). The fifth and seventh building blocks described in this study, namely the Error Correction Module and Active Learning Module, provide this functionality. We expect that algorithm errors will be highest when the algorithm is presented with input data that occurred in low frequency during training. These modules provide mechanisms for a radiologist to identify these underfit data to be submitted for algorithm retraining and improvement.

Although the particular implementation of the integration modules in this paper was aimed at addressing the needs of a particular use-case, the building blocks define generalizable tools needed for AI integration in a variety of radiology settings. A well-adopted, standards-based approach to these building blocks, and standards that govern the communications between these building blocks, will enable AI developers in the radiology space to more easily deploy their solutions in clinical environments.

Finally, radiology imaging data forms just one piece of the larger healthcare data puzzle, with spending on all AI-related healthcare tools estimated to top $8 billion by 2022 (20). These data must be integrated and discoverable within the larger healthcare information ecosystem.The technical aspects of integration of AI into healthcare must be supported by careful studies that prove its value beyond current techniques.

## CONCLUSION

Image analysis algorithms can enable radiologists to perform their jobs more accurately and efficiently. We describe seven building blocks that optimize the integration of these algorithms into clinical workflow. The implementation of these building blocks in this study can be used to inform development of more robust, standards-based solutions.

## Data Availability

N/A

